# Evaluating recovery, cost, and throughput of different concentration methods for SARS-CoV-2 wastewater-based epidemiology

**DOI:** 10.1101/2020.11.27.20238980

**Authors:** Zachary W. LaTurner, David M. Zong, Prashant Kalvapalle, Kiara Reyes Gamas, Austen Terwilliger, Tessa Crosby, Priyanka Ali, Vasanthi Avadhanula, Haroldo Hernandez Santos, Kyle Weesner, Loren Hopkins, Pedro A. Piedra, Anthony W. Maresso, Lauren B. Stadler

## Abstract

As the COVID-19 pandemic continues to affect communities across the globe, the need to contain the spread of the outbreaks is of paramount importance. Wastewater monitoring of the SARS-CoV-2 virus, the causative agent responsible for COVID-19, has emerged as a promising tool for health officials to anticipate outbreaks. As interest in wastewater monitoring continues to grow and municipalities begin to implement this approach, there is a need to further identify and evaluate methods used to concentrate SARS-CoV-2 virus RNA from wastewater samples. Here we evaluate the recovery, cost, and throughput of five different concentration methods for quantifying SARS-CoV-2 virus RNA in wastewater samples. We tested the five methods on six different wastewater samples. We also evaluated the use of a bovine coronavirus vaccine as a process control and pepper mild mottle virus as a normalization factor. Of the five methods we tested head-to-head, we found that HA filtration with bead beating performed the best in terms of sensitivity and cost. This evaluation can serve as a guide for laboratories establishing a protocol to perform wastewater monitoring of SARS-CoV-2.

**Highlights:**

- Five methods for concentrating SARS-CoV-2 RNA from wastewater evaluated
- Method performance characterized via recovery, cost, throughput, and variability
- HA filtration with bead beating had highest recovery for comparatively low cost
- Bovine coronavirus, pepper mild mottle virus assessed as possible recovery controls

## 1. Introduction

As the COVID-19 pandemic impacts millions worldwide, community monitoring and early detection of disease outbreaks has become of critical importance. While the rate of clinical testing has seen dramatic global increases since the pandemic onset, difficulties in assessing community health via this method remain (Tromberg et al., 2020). These difficulties include the logistics and cost of clinical testing, and a lack of robust contact tracing protocols in most communities. Moreover, the opt-in nature of clinical testing means asymptomatic and symptomatic individuals who decide to forego testing are not accounted for in community prevalence estimates.

Wastewater-based epidemiology (WBE) is an emerging paradigm for monitoring the community prevalence of SARS-CoV-2 (Hart and Halden, 2020). WBE has already proven to be a viable method for community monitoring of other viral pathogens, including poliovirus, rotavirus, hepatitis A virus, hepatitis E virus, noroviruses, enteroviruses, and adenoviruses, suggesting it could be appropriate for SARS-CoV-2 surveillance (Hellmér et al., 2014; Kamel et al., 2011; Katayama et al., 2008; Lago et al., 2003; McCall et al., 2020). This method of monitoring for SARS-CoV-2 is possible due to fecal shedding of SARS-CoV-2 virus particles and/or virus RNA before, during, and after clinical symptoms manifest in infected individuals (Cheung et al., 2020; Mesoraca et al., 2020; Wölfel et al., 2020). Once feces containing SARS-CoV-2 RNA enters the sewershed, the viral RNA is transported to wastewater treatment plants (WWTP) where it can be detected and quantified.

The detection of SARS-CoV-2 RNA in untreated domestic wastewater has already been reported in numerous studies from countries across the globe including Australia, Italy, the United States, Japan, and more (Ahmed et al., 2020a; La Rosa et al., 2021; Sherchan et al., 2020; Torii et al., 2020). Moreover, concentrations of viral RNA have been shown to correlate with community prevalence of SARS-CoV-2 (Stadler et al., 2020). WBE alleviates roadblocks associated with clinical testing by providing a cheaper, less logistically challenging method for monitoring communities. Importantly, it does not require individuals to opt-in, thereby capturing both symptomatic and asymptomatic individuals. The potential of WBE to inform public health measures by offering trend tracking and prevalence estimates is already of growing interest to local governments and has been put to use by universities to proactively prevent COVID-19 outbreaks in campus housing (Colosi et al., 2020; Stadler et al., 2020).

The development of rapid, cost-effective, and sensitive methods for quantifying SARS-CoV-2 RNA in wastewater are essential for widespread, successful implementation of WBE for SARS-CoV-2. Broadly speaking, the detection and quantification of viral RNA in wastewater is achieved through four steps: (1) wastewater sampling, (2) wastewater concentration, (3) RNA extraction, and (4) RNA quantification. There are multiple methods to choose from for each step, with disparate effects on the performance and practicality of the overall measurement system. As of now, there are no standard or clearly optimal methods for each step and methods are often selected based on a review of the literature, author familiarity with the method, and equipment and/or budget. Therefore, there is a need for the SARS-CoV-2 WBE community to better characterize and directly compare different methods for quantifying SARS-CoV-2 RNA in wastewater.

In this work we characterized different methods for concentrating SARS-CoV-2 RNA from wastewater. We focused on the concentration step because wastewater concentration methods applied for SARS-CoV-2 RNA vary widely from electronegative filtration with bead beating (Ahmed et al., 2020a), electronegative filtration with elution (Sherchan et al., 2020), ultrafiltration (Westhaus et al., 2021), precipitation (La Rosa et al., 2021), ultracentrifugation (Prado et al., 2020), and direct extraction (Crits-Christoph et al., 2020). Moreover, differences in approach, such as sample volume or whether to separate solids by centrifugation can impact measurement outcomes. Without standardization, it has been difficult to compare concentration methods across sites, study their relative strengths and weaknesses, optimize the methods, and understand the biggest sources of RNA loss. Despite these obstacles, our recent study had success in applying empirical adjustment factors to account for differences in SARS-CoV-2 RNA measurement methods between two labs (Stadler et al., 2020). The lack of internal standards presents another caveat, as RNA recovery percentages from wastewater samples vary between methods and thus non-normalized viral concentrations may impact comparisons across sites. In order to address this problem, surrogate viruses can be used to estimate the recovery efficiency and quantification of a target virus. Recent studies have used surrogate viruses to estimate SARS-CoV-2 RNA recovery percentages across a variety of methods (Ahmed et al., 2020a; La Rosa et al., 2021; Sherchan et al., 2020; Torii et al., 2020). To our knowledge, only one study has compared SARS-CoV-2 recovery from wastewater across methods in depth, but there were significant differences between compared methods that make it difficult to pinpoint the concentration step or some other factor as the source of differences (Pecson et al., 2020). At least one review compared concentration methods across studies, but there is again other differences between studies that make it difficult to directly compare the concentration method (La Rosa et al., 2020)

In this study, we performed a head-to-head comparison of five different concentration methods on samples from six wastewater treatment plants (WWTPs) in Houston, TX. We evaluated the methods by comparing the yields of SARS-CoV-2 RNA and sensitivity of detection by each method. In addition, we compared the recovery of a spiked surrogate virus, bovine coronavirus (BCoV), and the yield of a fecal indicator virus, pepper mild mottle virus (pMMoV). We provide a comprehensive practical summary for each concentration method by outlining start-up cost, consumable cost per sample, throughput time, limit of quantification (LoQ), and the variation in N1 and N2 detection between replicate samples. Overall, this study involves an extensive analysis of concentration methods currently in use to quantify SARS-CoV-2 RNA in wastewater, and an examination of BCoV and pMMoV as potential control factors.

## 2. Methods

### 2.1 Surrogate Preparation

BCoV was chosen as a surrogate and quality control measure to analyze viral RNA recovery between different concentration methods. Calf Guard (Zoetis) cattle vaccine containing an attenuated strain of the surrogate was used as the source for BCoV. Freeze-dried virus in 3 mL vials was rehydrated in sterile conditions with 1.5 mL of TE buffer on the morning of sample collection. Multiple vials were rehydrated and combined to prepare enough stock solution to spike all samples. A 100 μL aliquot of BCoV was immediately stored at −80L to later determine the concentration of BCoV in the stock solution.

### 2.2 Wastewater Sampling

Time-weighted composite samples of raw wastewater (influent) were collected every 1 hour for 24 hours. The collection period began the morning of Monday, October 5, 2020 and ended the morning of Tuesday, October 6, 2020. Wastewater samples from 6 facilities with a range of compositions, as measured by total suspended solids (TSS), carbonaceous biochemical oxygen demand (CBOD), and ammonia (NH4-N), were collected to test the robustness of the different concentration methods **(Table 1)**.

**Table 1.** Characteristics of the different wastewater treatment systems. Average daily flow rate was recorded on the day of October 5. Population data was extracted from the 2019 American Community Survey (*U.S. Census Bureau, 2019*). Composition data were reported from samples taken between October 5, 2020 to October 6, 2020. All data was provided by the City of Houston (Houston Public Works and Houston Health Department).

| Wastewater Treatment Plant (Anonymized) | Average Daily Flow Rate (MGD) | Population | TSS (mg/L) | CBOD (mg/L) | NH<sub>4</sub>-N (mg-N/L) |
| --- | --- | --- | --- | --- | --- |
| A | 8.52 | 167,000 | 342 | 126 | 26.2 |
| B | 16.6 | 304,000 | 970 | 120 | 17.4 |
| C | 9.69 | 330,000 | 196 | 155 | 26.2 |
| D | 0.180 | 13,400 | 100 | 155 | 34.2 |
| E | 3.21 | 86,600 | 58.7 | 108 | 30.5 |
| F | 1.53 | 48,200 | 106 | 201 | 32.0 |

### 2.3 Wastewater Sample Collection

At the end of the sampling period, all samples were transported on ice to a central processing facility. Larger sample volumes were aliquoted into 500 mL, Nalgene™ Wide-Mouth HDPE Packaging Bottles (3121890016, Thermo Scientific) and spiked with 50 μL of BCoV stock solution. Aliquots were transported on ice to Baylor College of Medicine or Rice University and then stored at 4°C for further processing.

### 2.4 Wastewater Sample Concentration

The different concentration methods are depicted in **Figure 1**. Concentration occurred the day following sample collection (Oct. 7). PEG concentration began the day of sample collection to allow the samples to sit overnight (Oct. 6). Technical replicates were performed in triplicate for each concentration method. The direct extraction, HA filtration with bead beating, and ultrafiltration concentration methods were completed at Rice University. The resulting concentrates were immediately transported to Baylor College of Medicine on ice for extraction. HA filtration with elution and PEG methods were completed at Baylor College of Medicine. Concentration methods were split between labs to reduce processing burden and because each lab had more experience with their respective methods. Concentrates were stored at 4°C until extraction.

**Figure 1:**
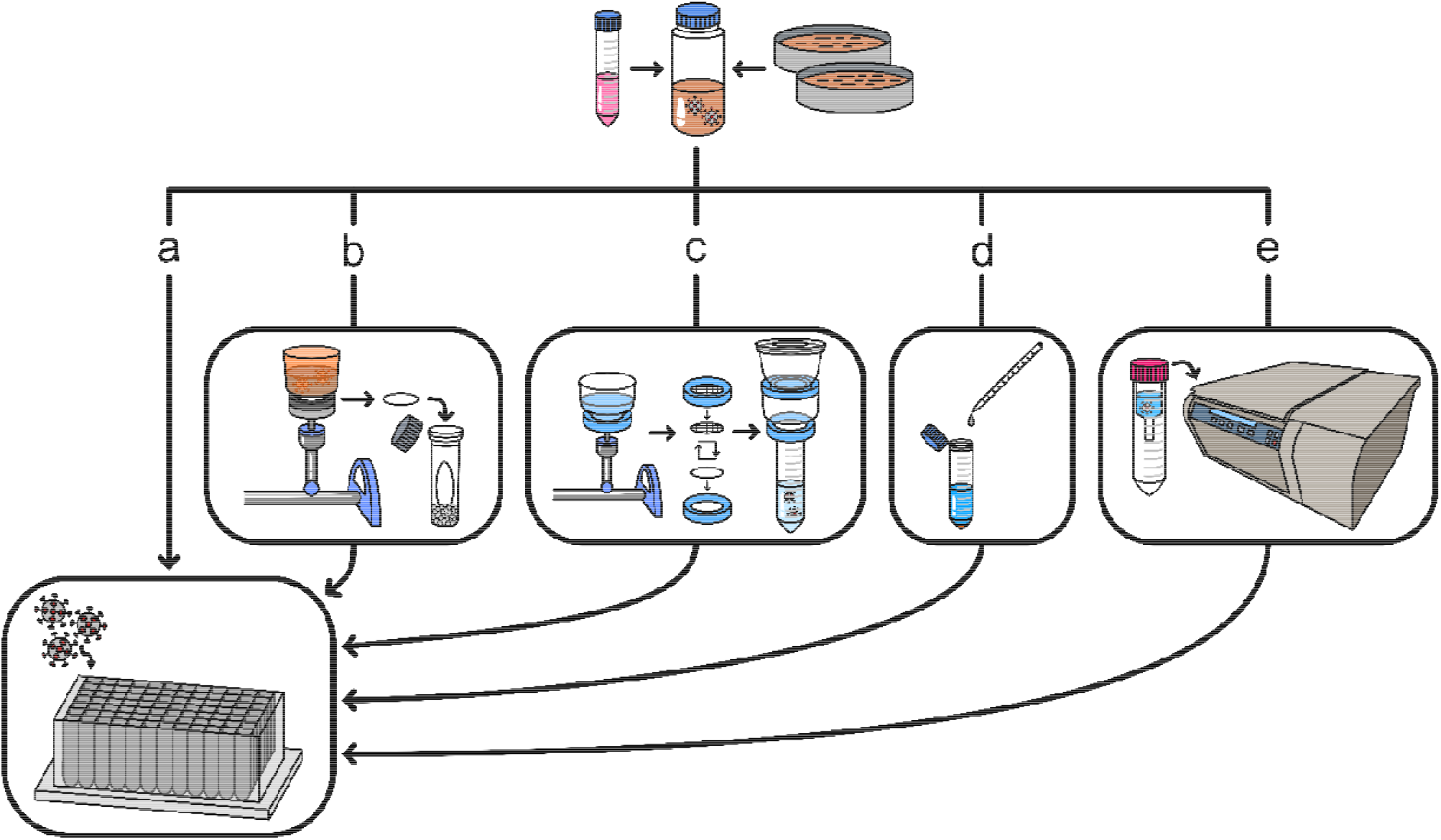
Overview of the evaluated concentration methods. Wastewater samples were collected from several wastewater treatment plants across Houston in sample collection bottles and immediately spiked with BCoV (top). a-e): The samples were then concentrated through several methods: a) direct extraction, b) HA filtration with bead beating, c) HA filtration with elution, d) PEG precipitation, and e) ultrafiltration. All concentrated samples subsequently underwent RNA extraction. Samples undergoing direct extraction were not concentrated and instead were directly extracted from the liquid phase of the wastewater samples.

#### 2.4.1 Direct Extraction

Approximately 1 mL of sample was aliquoted into a 1.5 mL centrifuge tube. The sample was then centrifuged for 5 minutes at 17,000 g and 4°C. The supernatant was carefully aspirated without disturbing the pellet and used for extraction.

#### 2.4.2 HA Filtration with Bead Beating

Roughly 50 mL of each sample was aliquoted into 50 mL conical tubes (1184R09, Thomas Scientific) and then centrifuged for 10 minutes at 4,100 g and 4°C. Prior to sample addition, the assembled MF 3, 300ml Magnetic Filter Holder with lid kit (200300-01, Sterlitech) was attached to the Multi-Vac 600-MS Manifold (180600-01, Sterlitech) and Rocker 800 Oil Free Laboratory Vacuum Pump (167800, Sterlitech) system. Electronegative Microbiological Analysis Membrane HA Filters (HAWG047S6, Millipore Sigma) were placed into the manifold system with sterile forceps prior to the filtration process. The filters were then washed with approximately 50 mL of ultrapure water before sample addition. A graduated cylinder was used to measure 50 mL of supernatant which was then poured directly into the assembled filter holder. After sample was added into the manifold system, 1 mL of 1.25 M MgCl_2_•H_2_O (M0250-500G, Sigma Aldrich) was added directly to the sample to achieve a final concentration of 25 mM. The samples were then gently swirled with a pipette tip to homogenize and allowed to sit for five minutes. The vacuum pump was subsequently turned on and allowed to pull the sample through the filter. After the sample passed through the filter, the vacuum pumps were turned off, the filters were rolled up with sterile forceps, and then placed into a filled bead beating tube (0.1 mm diameter glass beads, Ca. No.: 11079101, BioSpec. Bead beating tube, Ca. No: 02-682-558, Fisher Scientific).

#### 2.4.3 HA Filtration with Elution

The beginning of the HA filtration with elution method was similar to HA filtration with bead beating. However, centrifugation for the elution method was completed for 1 minute at 3,000 g and 4°C. Furthermore, the use of EZ-Fit™ Filtration Unit (EFHAW100B, Millipore Sigma) is unique to the elution method. These are sterile, single use filter holders that come with the same electronegative filters used in HA filtration with bead beating. These units were used to facilitate the elution aspect of the elution method. The major divergence from HA filtration with bead beating is how the captured virus was recovered from the filters after filtration. After filtration, filters were carefully flipped over with sterile forceps and placed back into the filter holder. Next, 5 mL of 1 mM NaOH (S318-100, Fisher Scientific) eluent was placed on top of the inverted filter. The back of a EZ-Fit Filtration Unit was then used to push the eluent through the filter and into a 15 mL conical tube (1184R08, Thomas Scientific) containing 12.5 μL of 100 mM H_2_SO_4_ (A300-212, Fisher Scientific) to neutralize the NaOH. Roughly 2.5 mL of eluent was collected from the setup. For concentration calculation purposes, the virus was treated as eluting into 2.5 mL.

#### 2.4.4 PEG precipitation

Solids were first removed by centrifuging wastewater in 500 mL centrifuge bottles (47735-696, VWR) for 15 minutes at 7,140 g and 4°C. The supernatant was then filtered through 0.22 μm Steritop Threaded Bottle Top Filters (SCGPS05RE, Millipore) into glass bottles. Then, 200 mL of sample was transferred into a new sterile 500 mL bottle. Next, 16 g of PEG 8000 (8% w/v) (VWRV0159-1KG, VWR) and 5.844 g of NaCl (0.5 M) (S271-1, Fisher Chemical) were added to the bottle. The solution was then inverted, gently shaken by hand, and allowed to precipitate overnight at 4°C. The following day, the sample was centrifuged for 30 minutes at 16,900 g and 4°C. The supernatant was then poured off and the pellet was resuspended in 2 mL of 1X PBS solution (0.01 M). The 1X PBS solution (0.01 M) was prepared with 1.096 g Na_2_HPO_4_ (S375-500, Fisher Chemical), 0.3148 g H_2_PO_4_Na•H_2_O (S369-500, Fisher Chemical), and 8.5 g NaCl (BP358-10, Fisher Chemical) per liter in ultrapure water and subsequently passed through a 0.22 μM filter into a sterile container. 1 mL of each suspension was then aliquoted into 1.5mL microcentrifuge tubes.

#### 2.4.5 Ultrafiltration

Roughly 50 mL of each sample was aliquoted into 50 mL conical tubes. The conical tubes were then centrifuged for 10 minutes at 4,100 g and 4°C. 50 mL of solids-free supernatant was then transferred to another 50 mL conical tube and stored on ice until further processing. The Amicon® Pro Purification System with 100kDa Amicon® Ultra-0.5 Devices (ACS510024, Millipore Sigma) were filled with 15 mL of ultrapure water and centrifuged for 8 minutes at 1,900 g and 4°C. Flow through and residual concentrate were poured out. Approximately 15 mL of supernatant was then loaded into each ultrafiltration device. The ultrafiltration devices were then centrifuged for 10 minutes at 4,100 g and 4°C. Flow through was discarded, and additional sample added to reach the volume limit of the ultrafiltration devices. The centrifugation and discarding process was repeated until each sample had completely passed through the ultrafiltration devices. As the ultrafiltration approached completion, centrifugation intervals of 3 minutes and 5 minutes were used to end with a final concentrate volume of approximately 1.5 mL. Concentrate was pipetted from the ultrafiltration devices into 2 mL microcentrifuge tubes. Microcentrifuge tubes containing the sample were weighed to calculate total volume of concentrate.

### 2.5 RNA Extraction

Liquid samples were extracted with the chemagic™ Prime Viral DNA/RNA 300 Kit H96 (CMG-1433, PerkinElmer) following manufacturer protocol. 300 μL of liquid concentrate was extracted into 100 μL of sterile, nuclease free water. Extraction occurred the day following concentration for all liquid samples.

Bead beating tubes containing filters from the HA Filtration with bead beating concentration method were extracted by first adding 600 μL of lysis buffer from the chemagic kit. The samples were then bead beaten on a FastPrep-24™ 5G bead beater (116005500, MP Biomedical) for 1 minute at 5 m/s, placed on ice for 2 minutes, bead beaten for 1 minute at 5 m/s, and then placed on ice. Samples were then centrifuged for 3 minutes at 17,000 g and 4°C. 300 μL of supernatant was then removed and subjected to the same chemagic extraction as used on the liquid samples. RNA extracts were stored at −80°C for 10 days and then transferred to −20°C for two days until quantification.

### 2.6 Quantification

#### 2.6.1 Reverse Transcription - Droplet Digital Polymerase Chain Reaction (RT-ddPCR)

One step RT-ddPCR was conducted with One-Step RT-ddPCR Advanced Kit for Probes (1864021, Bio-Rad) on the QX200 AutoDG Droplet Digital PCR System (Bio-Rad) to quantify the concentration of N1 SARS-CoV-2, N2 SARS-CoV-2, and M BCoV gene targets in extracted samples. Primer and probe information can be found in **Table S1**. Reaction mixes were prepared on ice according to the composition outlined in **Table S2** for N1 and N2, and **Table S3** for BCoV. RNA template for BCoV was diluted 50x to attain a concentration within the quantifiable range of the ddPCR equipment. Thermocycling conditions are outlined in **Table S6**. After thermocycling, samples were held at 4°C for no longer than 12 hours until being read on the QX200 Droplet Reader (18644003, Bio-Rad). Droplet data was analyzed on the QuantaSoft v1.7.4 software. Manual thresholding of droplets was only performed when QuantaSoft was unable to automatically threshold.

#### 2.6.2 Reverse Transcription - Quantitative Polymerase Chain Reaction (RT-qPCR)

One step RT-qPCR was conducted with qPCRBIO Probe 1-Step Go Separate-ROX (PB25.44-12, PCR Biosystems) on the QuantStudio 3 Real Time PCR System (A28567, Applied Biosystems) to quantify the concentration of VGP pMMoV gene targets in extracted samples. Primer and probe information can be found in **Table S1**. Reaction mixes were prepared on ice according to the composition outlined in **Table S4**. RNA template for pMMoV was taken from the 50x dilutions created for measuring BCoV to conserve undiluted extract. Thermocycling conditions are outlined in **Table S7**. qPCR data was analyzed on the QuantStudio Design and Analysis v1.4 software.

Standards of linear DNA (IDT), 708 bp in length, were prepared and ran in triplicate in a dilution series with concentrations at 0.69, 6.9, 69, 690, 6,900, 69,000, to 690,000 gene copies/µL standard. Herring sperm DNA (D1811, Promega) at a final concentration of 10 ng/µL was used to dilute the standards as a carrier DNA to preserve pMMoV standard DNA fragments during freeze-thaws.

#### 2.6.3 Limit of Quantification (LoQ)

The LoQ for ddPCR was defined as 3 positive droplets per 10,000 total droplets generated by the instrument as recommended by the manufacturer. The volume of an individual droplet (0.86 nL) was then used to calculate a LoQ of 0.767 gene copies/µL RNA template for a reaction setup with 10 µL of RNA template.

The LoQ for RT-qPCR was determined to be 0.69 gene copies/µL RNA template for a reaction setup with 4 µL of RNA template. This was the concentration of the lowest standard used in our calibration curve. pMMoV was the only gene target measured through RT-qPCR, and all values were significantly above this limit. Concentration factors **(Table S8-S13)**, average percent recovery, and unit conversions were then used to convert this raw LoQ to an effective LoQ associated with each concentration method **(Eqn. 1)**.

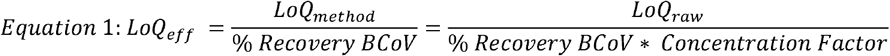

Measurements below the LoQ may indicate presence of gene targets but are not reliably accurate measurements of the concentration.

## 3. Results & Discussion

### 3.1 Quantification of SARS-CoV-2 RNA in Wastewater with Different Concentration Methods

We first evaluated the effectiveness of five concentration methods at detecting and quantifying the SARS-CoV-2 RNA in wastewater samples. We obtained 24 hour composite wastewater samples from six different WWTPs in Houston covering a range of influent flow rates, population sizes, and wastewater compositions **(Table 1)**. We also created a negative control sample that contains only DI water that had been spiked with the BCoV surrogate. We applied each of the five different concentration methods to each sample in triplicate, extracted the RNA of the SARS-CoV-2 virus, and quantified the concentration of CDC target N1 and N2 using digital droplet PCR (ddPCR). We then back calculated the concentration of the virus in terms of copies of the virus per liter of wastewater **(Figure 2A, B)**.

**Figure 2.**
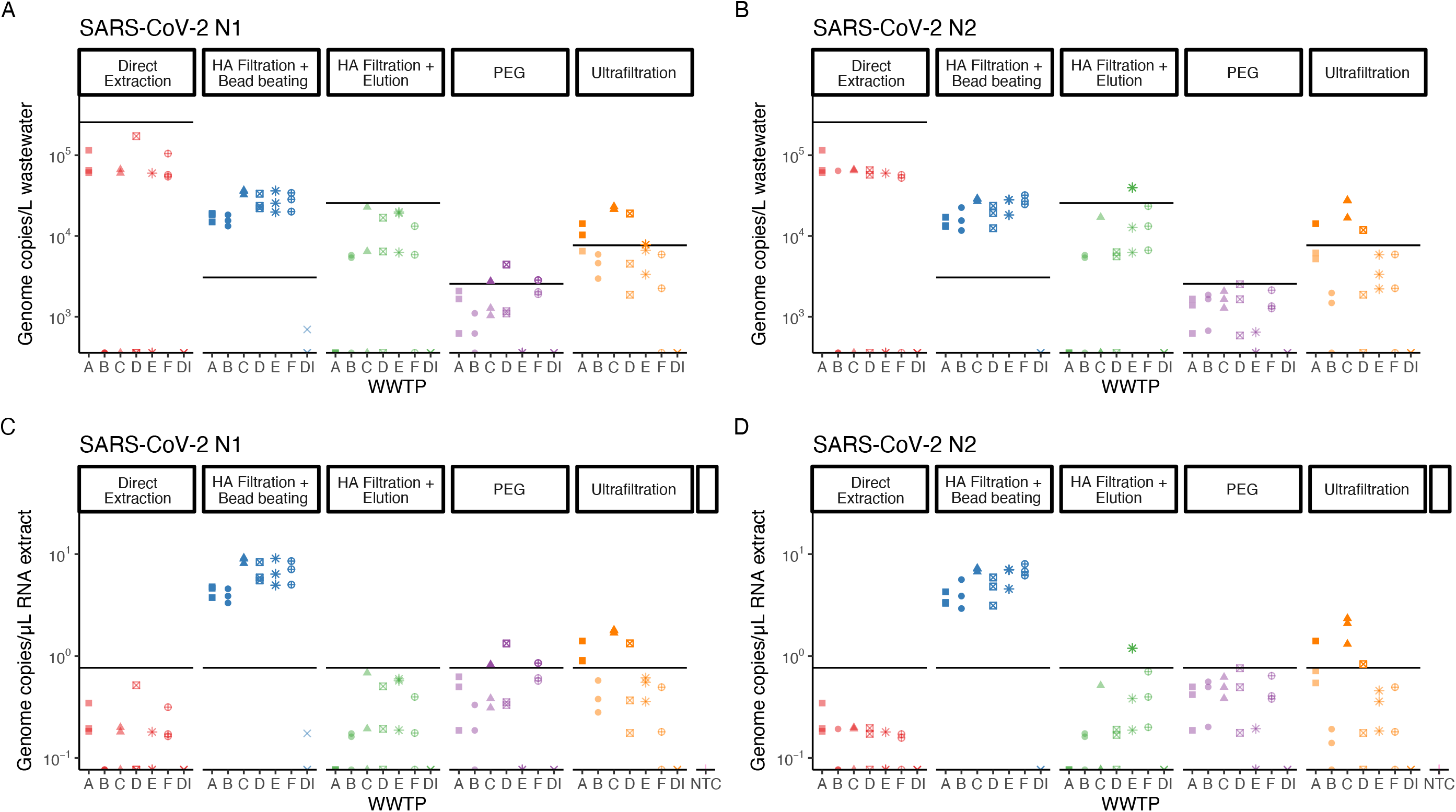
SARS-CoV-2 RNA Concentrations. N1 (a) and N2 (b) gene target concentrations determined using different concentration methods for six wastewater samples reported in gene copies/L wastewater. N1 (c) and N2 (d) gene target concentrations determined using different concentration methods for six wastewater samples reported in gene copies/uL RNA template. Black horizontal lines indicate LoQs. WWTP are A-E, DI is deionized water, and NTC is no template control.

The concentration methods use different mechanisms to concentrate SARS-CoV-2. HA filtration concentrates via manipulation of charge interactions between virus particles and filter media (Cashdollar and Wymer, 2013). The addition of salts effectively replaces the repulsive interactions between the negatively charged virus surface and the negatively charged surface of the filter with positive-ion bridges. The bead beating method then desorbs and ruptures virus particles absorbed to the filter membrane surface. In the HA filtration with elution method, an eluent is added that desorbs the virus particles from the filter membrane surface into a smaller volume by altering the pH. The PEG method concentrates by precipitation of virus particles upon addition of polyethylene glycol and sodium chloride. Although there is uncertainty in the exact mechanism, virus precipitation is believed to occur similarly to precipitation of proteins by PEG, where water molecules are drawn from the solution to hydrate PEG molecules, thereby increasing the effective protein concentration, making it insoluble, enabling the proteins to precipitate after reaching saturation (Ingham, 1990; Yamamoto et al., 1970). Ultrafiltration concentrates via size exclusion, allowing water and other small particles to pass through a filter, but blocking larger sized particles like SARS-CoV-2 virus (Cashdollar and Wymer, 2013). The different mechanisms involved in these concentration methods lead to different degrees of recovery of SARS-CoV-2, BCoV, and pMMoV.

All five of the tested concentration methods were able to detect the presence of SARS-CoV-2 RNA, but varied significantly in viral RNA titer. Direct extraction yielded the highest apparent concentration of SARS-CoV-2 RNA across all wastewater samples. In contrast, PEG had consistently lower signal for SARS-CoV-2 RNA. Both methods involving HA filtration (with bead beating and with elution) yielded similar concentrations that were about a half log lower than direct extraction. In all cases, the resulting concentration of N1 and N2 per liter of wastewater was highly dependent on the concentration method used, more so than from which WWTP the sample came from. This suggests that the true concentration of SARS-CoV-2 RNA from all six WWTPs was roughly the same. It is worth noting that these measurement systems may be detecting free SARS-CoV-2 RNA along with intact SARS-CoV-2 virus particles, and that each concentration method may have a different ability to measure that free SARS-CoV-2 RNA.

Direct extraction yielded the highest concentrations of genome copies per L of wastewater due to the fewest losses associated with concentration and the largest concentration factor applied. When the results are shown in terms of the raw data, copies of N1 or N2 per µL of RNA template, direct extraction had the lowest raw concentrations of viral RNA and many data points were below the LoQ (as shown by the line in (**Figure 2C, D**). Furthermore, the only method that yielded results that were consistently above the LoQ for all WWTPs and both targets was HA filtration with bead beating. Because the LoQ is constant in terms of copies of RNA per µL of RNA template, the method that yields the highest raw concentration of RNA per µL of template is the one that produces quantifiable signal. Therefore, when evaluating concentration methods, a key metric for consideration is not the genome copies per liter wastewater, but the copies per µL of RNA template. In the set of methods we evaluated, we found that HA filtration with bead beating performed the best in terms of having the highest raw genome copies per microliter of RNA template and was thus consistently able to quantify SARS-CoV-2 RNA in all wastewater samples tested.

One important decision in SARS-CoV-2 RNA concentration that may impact sensitivity, reproducibility, and variability is the volume of input wastewater. As input volume increases you are left with a higher concentration of virus RNA in a concentrate of the same volume. However, this does not come without sacrifice. In all three filtration-based methods, there is a nearly exponential relationship between input volume and processing time (data not shown), which can be an issue for the logistics of WBE for SARS-CoV-2. There is also an upper limit on filterable volume, as pores in the filter membrane become completely blocked. In the case of PEG, larger input volume means larger volumes need to be placed in centrifuges which have maximum volumetric capacities and increases in startup price associated with increasing that capacity by purchasing higher-powered centrifuges. Additionally, the fraction of recovered virus may not stay constant for all methods with a higher volume. In the case of HA filtration methods, for example, absorption sites on the filter surface may be increasingly occupied as additional volume is filtered, reducing the amount of virus particles and virus RNA that can absorb to the surface. All concentration methods in this study, except for direct extraction and HA filtration with elution, used 50 mL of input volume. Direct extraction used 300 μL due to volume limits of the extraction kit that we used, and HA filtration with elution used 25 mL due to rate of filtration limits in the operating lab. An in-depth investigation into volume optimization was out of the scope of this study but could be performed to improve the sensitivity and reduce the variability of each of the concentration methods.

### 3.2 Practical considerations of different concentration methods

Next, we compared characteristics related to the practicality of the different concentration methods. We did this comparison to help relevant parties decide which SARS-CoV-2 RNA concentration method is best suited for their situation. Laboratories might have differences in availability of resources (equipment, labor) and in the number of samples requiring analysis, and thus can use these results to optimize workflow.

The startup costs reflect equipment that was unique to each concentration method. Standard lab equipment required by all concentration methods, such as pipettes and PPE, were not included in these calculations. Additionally, RNA extraction costs were not included, except in the case for the price of a bead beater, which is necessary to quantify RNA when using the HA filtration with bead beating method. The largest factor contributing to startup costs in all cases was centrifuges. The differences in startup costs was generally due to differences in cost of the specific centrifuge required. A centrifuge is not necessary to perform the HA filtration with bead beating method, but centrifuging samples prior to concentration to remove solids drastically decreased filtering time and thus greatly increased throughput efficiency (data not shown). Pecson et al. also found that a solids removal step did not show a clear impact on the quantification results of SARS-CoV-2 (Pecson et al., 2020). A detailed breakdown of startup costs can be found in **Table S14-S18**.

By far the most expensive method in terms of consumables cost per sample is ultrafiltration. The Amicon® filters required by this process are expensive and not reusable. The PEG method is also relatively high in cost for a similar reason; the unique bottom top filters are costly and cannot be reused. It may be possible to lower costs for PEG by purchasing individual 0.22 µm filters to use in a filter manifold setup, however, this would further sacrifice throughput, as time would be required to rinse the manifold setup to prevent cross contamination. HA filtration with bead beating and elution methods had comparable consumables costs, although the elution method is more expensive due to the use of an EZ-Fit™ Filtration Unit. We do not suggest reusing or replacing the EZ-Fit units to lower cost, since contact with the flipped filter and underlying surface poses a large cross contamination risk if done in a repeatedly used filter manifold. Direct extraction was the least expensive method since specialized consumables were not required. A detailed breakdown of consumable costs can be found in **Table S19-S23**.

Throughput time was also assessed as different groups conducting WBE for SARS-CoV-2 have varying numbers of samples and unique requirements for turnaround times needed to report data. All concentration methods included a centrifugation step to remove solids in their throughput time. The PEG method had the lowest throughput, largely because of the precipitation step, taking at least 3 hours longer to process a batch of samples than the other methods evaluated. In our study, PEG was precipitated overnight to lower the burden on lab workers, but our experience with PEG concentration suggests that this time could be lowered to 4 hours without lowering recovery. Other literature concentrating viruses using PEG have reduced precipitation time further, but investigation of the effect of this on recovery was out of the scope of this study. Ultrafiltration had the second lowest throughput due to non-filterable matter accumulation during centrifugation. In centrifugation steps that allow fluid to pass directly through a filter, clogging and precipitation is probable, thus increasing processing time. Direct extraction had the highest throughput efficiency, since it only required a five minute solids removal step. HA filtration with bead beating and HA filtration with elution had high and comparable throughputs. A detailed breakdown of throughput can be found in **Table S24**.

We then determined the LoQ for all of the concentration methods. Here we defined the LoQ by translating the minimum droplet counts required to reliably quantify gene targets (an approach recommended by Bio-Rad). The minimum droplet count, 3 positive droplets per 10,000 total droplets, translates to 0.767 gene copies/μL ddPCR reaction. The amount of RNA template and different concentration factors between methods were then used to calculate the LoQ in **Table 2**. PEG had the highest concentration factor of 300, leading to the lowest LoQ, while direct extraction has the highest LoQ because there was no concentration occurring. It is important to note that this LoQ does not take into account losses incurred by the different concentration methods. The optimal method will balance a low LoQ with a high recovery leading to raw concentrations of gene targets being significantly above the raw LoQ. We accounted for recovery by averaging percent recovery of BCoV for the different methods and incorporating that average into an effective LoQ. It should be noted that BCoV has yet to be identified as an optimal surrogate for SARS-CoV-2 but can still provide valuable information in this context. The recovery of BCoV through PEG, averaging 0.09%, was low enough to increase the effective LoQ to higher than that of HA filtration with bead beating. When incorporating percent recovery of the different methods, HA filtration with bead beating had the lowest effective LoQ at 2.76e5 gene copies/L wastewater, and was almost an order of magnitude lower (more sensitive) than the other concentration methods **[Table 2]**. The effective LoQ is directly related to the sensitivity of the concentration method, which is a critical factor in being able to reliably quantify SARS-CoV-2 in wastewater samples, especially when there is relatively low community prevalence. Lowering the effective LoQ relative to direct extraction is the essential reason why we include a concentration step, because it makes lower concentrations of virus more reliably quantifiable.

**Table 2:** Summary of key metrics of each concentration method. A detailed breakdown of costs and list of equipment used can be found in S14-23.

| Method | Startup cost (\$) | Consumables cost (\$/ sample) | Throughput (hr/6 samples) | LoQ (gc/L WW) | Effective LoQ (gc/L WW) | BCoV %CV | pMMoV %CV |
| --- | --- | --- | --- | --- | --- | --- | --- |
| **Direct extraction** | \$5,650 | \$0.14 | 0.1 | 2.56E+05 | 8.39E+06 | 14.6 | 29.9 |
| **HA Filtration + Bead Beating** | \$15,368 | \$1.50 | 0.7 | 3.07E+03 | 2.76E+05 | 27.3 | 25.9 |
| **HA Filtration + Elution** | \$11,160 | \$5.80 | 0.5 | 2.56E+04 | 3.89E+06 | 20.9 | 31.8 |
| **PEG** | \$20,288 | \$11.02 | 4.6 | 2.56E+03 | 2.70E+06 | 39.0 | 49.8 |
| **Ultrafiltration** | \$9,000 | \$12.10 | 1.5 | 7.67E+03 | 2.63E+06 | 24.4 | 49.5 |

In a study that investigated different overall processing methods for measurement of SARS-CoV-2, no systematic impact by the concentration method was found for results corrected with a process control (Pecson et al., 2020). As mentioned previously however, there were confounding factors, such as differences in extraction and quantification steps, between the different processing methods that could have skewed the comparison of the concentration step. Another study investigated the recovery of murine hepatitis virus (MHV), a proposed process control for SARS-CoV-2, between multiple concentration methods with the same extraction and quantification steps (Ahmed et al., 2020b). The general ranking of the recovery results for the concentration methods they tested generally agree with ours in the order of best to worst: HA filtration with bead beating, ultrafiltration, and PEG. A third study compared concentration of MHV via PEG, ultracentrifugation, and ultrafiltration and found that ultrafiltration was the preferrable concentration method for enveloped viruses of those three methods, but this study based its results on the quantification of live virus and not virus RNA (Ye et al., 2016). They suggest that PEG is a preferred method only for non-enveloped viruses. Overall, our study our study controlled for different confounding factors and directly measured SARS-CoV-2 RNA, unlike these previous two studies.

### 3.3 Recovery of corrective surrogates

In addition to SARS-CoV-2 RNA, we compared the concentrations of BCoV and pMMoV RNA in the WWTP samples processed via the different concentration methods. Each wastewater sample was immediately spiked with a known concentration of BCoV upon reception at the collection site. Samples that had passed through concentration and extraction protocols were then analyzed through ddPCR (BCoV) and qPCR (pMMoV) to determine their concentrations in the wastewater samples.

### 3.4 Bovine Coronavirus (BCoV) Process Control

A process control is necessary when exact RNA recovery efficiency of the target component across different processing steps is unknown, or when determining these recoveries may be impractical. By spiking in a known concentration of the process control at the beginning of the process or at different stages throughout the process and comparing the measured concentration to the expected concentration, it is possible to determine the overall recovery of the process control and loss of the process control during sample processing. If the process control has been validated, and is known to behave in a way similar to the target component, then these losses can be incorporated into the measured concentration of the target component to estimate a “true” measure of the target component in the sample. Process controls can also be used simply as positive controls to ensure that nothing went awry during sample processing and analysis.

Recent SARS-CoV-2 WBE studies have used a variety of process controls, including MHV (Ahmed et al., 2020b), transmissible gastroenteritis virus (TGEV) (Mlejnkova et al., 2020), human coronavirus (HCoV 229E) (La Rosa et al., 2021), Phi 6 (Sherchan et al., 2020), bovine respiratory syncytial virus (BRSV) (Gonzalez et al., 2020), and BCoV (Gonzalez et al., 2020). However, no single process control has proven, as of yet, to be significantly more indicative of SARS-CoV-2 recovery than other process controls. In this work we sought to assess BCoV as a process control by comparing its recovery across different methods and wastewater samples against the yields of SARS-CoV-2 RNA.

We chose BCoV because of its similarity to SARS-CoV-2, as both viruses are part of the genus *Betacoronaviridae*. SARS-CoV-2 is an enveloped virus, generally spherical in shape with mild pleomorphism (60-140 nm diameter) and a 29.9 kb length genome (Zhu et al., 2020). BCoV is a pleomorphic (65-210 nm diameter), enveloped RNA virus with a 27 - 32 kb length genome (Saif, 2010). Both BCoV and SARS-CoV-2 carry a spike (S) glycoprotein on their envelope surface, while only BCoV carries an additional, large protein on its envelope surface known as hemagglutinin-esterase (HE) glycoprotein (Saif, 2010). Apart from structural similarity, BCoV is also easily obtainable in an attenuated form from a common cattle vaccine and poses a low health risk to humans.

The magnitude of recovery of BCoV reflected the magnitude of recovery of N1 and N2 across concentration methods. Like N1 and N2, the highest to lowest recovery of BCoV from the different concentration methods occurred in the order of direct extraction, HA filtration with bead beating, HA filtration with elution, ultrafiltration, and finally PEG. Overall, this suggests that BCoV could be a good surrogate for SARS-CoV-2 as relative recovery of BCoV across concentration methods mirrors the relative recoveries of SARS-CoV-2. However, there is some difficulty in directly comparing recovery of N1 and N2 to BCoV due to the number of N1 and N2 measurements that were below the LoQ for ddPCR. Additionally, it is not clear what the dominant forms (intact viral particle vs free RNA) of BCoV and SARS-CoV-2 are when they reach the concentration step of the measurement process and how this affects concentration. More research is needed to characterize the form of SARS-CoV-2 in wastewater and understand how the form impacts concentration to inform the choice of an appropriate surrogate.

Interestingly, DI water controls spiked with BCoV showed significantly different levels of recoveries compared to wastewater samples concentrated via the same method. One potential explanation for this could be rupture of the viral particles due to a large difference in osmotic pressures across viral capsid/envelope in DI water. Rupture may not occur in wastewater samples due to a significant amount of dissolved compounds reducing differences in osmotic pressure inside and outside of the viral particle. Thus, the disparate concentration methods may have different effects on ruptured versus unruptured virus.

### 3.5 Pepper Mild Mottle Virus (pMMoV) Normalization Factor

There are many factors that affect wastewater concentration of SARS-CoV-2 RNA between excretion in feces and quantification in the lab that potentially confound translation of concentration to community prevalence. While a process control, like BCoV, can be used to account for factors during the measurement process (i.e. between sampling and quantification), a normalization factor attempts to account for factors during the measurement process and additional upstream factors, like dilution in the sewer system.

pMMoV has been suggested as a promising normalization factor for SARS-CoV-2 (Wu et al., 2020). It is the most abundant RNA virus found in human feces due to its origin in peppers and pepper containing products and has previously been proposed as a water quality and fecal pollution indicator (Kitajima et al., 2018; Rosario et al., 2009). In theory, it is excreted in relatively consistent amounts in humans across a population and will travel alongside SARS-CoV-2 viral particles and viral RNA in the conveyance system, experiencing the same conditions. pMMoV is a rod-shaped (∼312 nm length), non-enveloped RNA virus with a 6.4 kb length genome in the *Tobamovirus* family (Kitajima et al., 2018). Due to a number of structural differences between pMMoV and SARS-CoV-2, pMMoV is better used as a fecal indicator than as a corrective process control.

Direct extraction showed the highest recovery of pMMoV in all the wastewater samples **(Figure 3)**. Three other methods, HA filtration with bead beating, HA filtration with elution, and ultrafiltration showed roughly equivalent recoveries of pMMoV. The PEG method had the lowest recoveries of pMMoV of all the concentration methods. All of the resulting pMMoV measurements, with the exception of two, were well above the LoQ.

**Figure 3.**
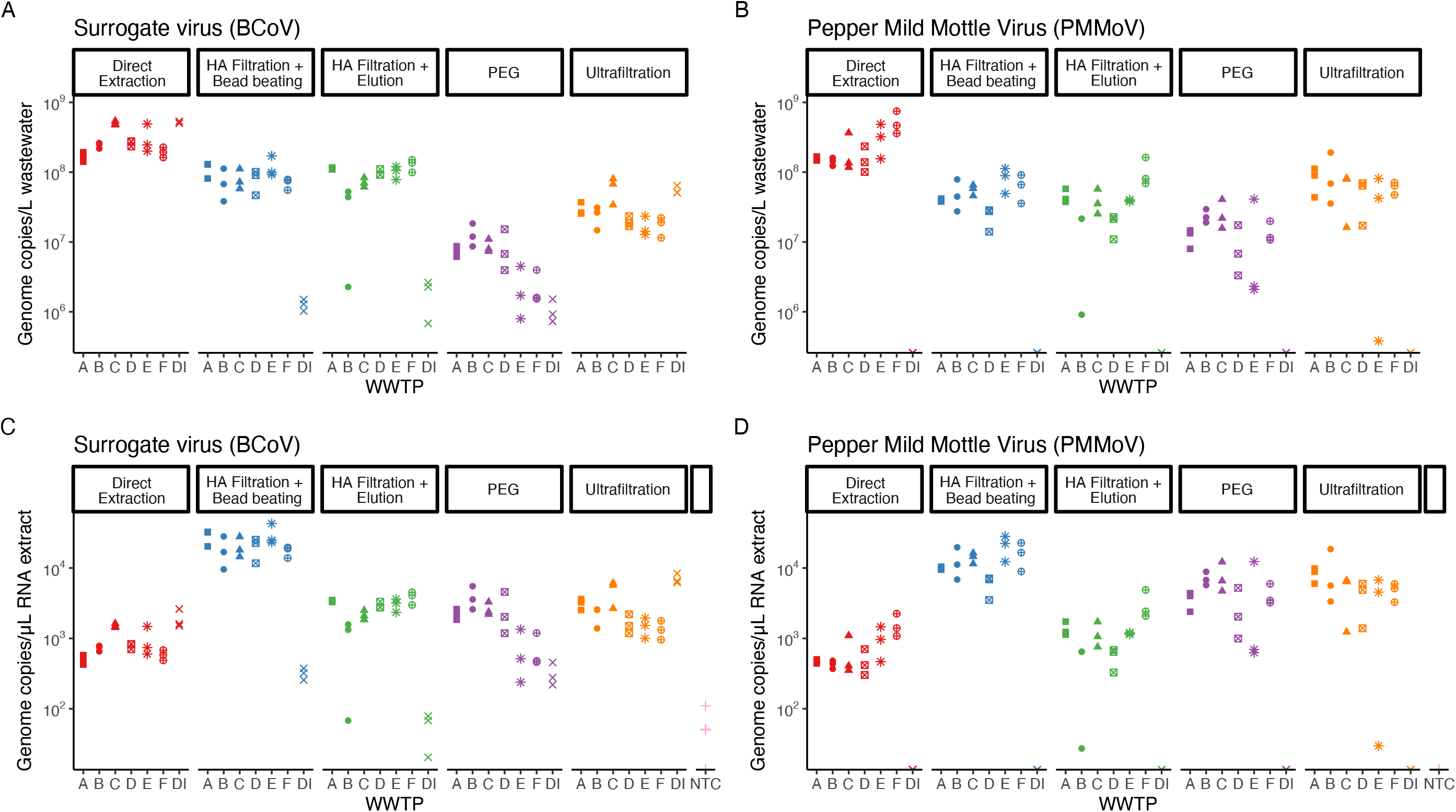
BCoV and pMMoV concentrations. Recovery of BCoV (a) and pMMoV (b) between different concentration methods and different WWTP reported in gene copies/L wastewater. Recovery of BCoV (c) and pMMoV (d) between different concentration methods and different WWTP reported in gene copies/μL RNA template. WWTP are A-E, DI is deionized water, and NTC is no template control.

The relative effectiveness of direct extraction and the PEG method compared to the other concentration methods was about the same between measurement of pMMoV and measurement of N1 and N2. However, while HA filtration with bead beating, HA filtration with elution, and ultrafiltration were roughly equivalent for pMMoV, HA filtration with bead beating recovered the most N1 and N2 of the three respective concentration methods.

A number of differences between pMMoV and SARS-CoV-2 may be the cause of these differences. For example, it may be that the protein capsid and envelope of SARS-CoV-2 are easier to rupture via bead beating than the sole protein capsid of pMMoV. Alternatively, it is possible that the forms of the virus are different after conveyance in the sewers system. One of the viruses may primarily exist in the form of free RNA due to decay of their envelope and/or protein capsid, while the other virus may be largely intact. It is currently not clear what form either pMMoV or SARS-CoV-2 are in when they reach the concentration step, how these forms impact concentration, or how different characteristics of the two viruses impact concentration, but they are areas that should be explored further.

### 3.6 Variability of measurement between concentration methods with BCoV and pMMoV

The variability of measurements between each concentration method was determined using the coefficient of variance (CV) for BCoV and pMMoV **(Table 2)**. The lower the CV, the lower the variability and the higher confidence one can have in a measured value. Further, if CV is low enough, it may be reasonable to reduce replicates (i.e. from triplicates to duplicates) to save on cost and throughput. The CV was measured for each WWTP for a particular method and then averaged for all WWTPs in the method to get the CV of the whole method. The PEG method showed the highest average CV, while the HA filtration with bead beating method had a generally low CV for both BCoV and pMMoV. As of now, it is not clear what causes the difference in variability between each concentration method. We chose not to directly incorporate CV of N1 and N2 gene targets because a large proportion of measurements were below the LoQ. Therefore variability in N1 and N2 would be more attributed to the quantification procedure that was used to establish the LoQ as opposed to variability caused by the particular concentration method. Between BCoV and pMMoV, the BCoV CV is likely a better indicator of a potential SARS-CoV-2 CV due to BCoVs higher structural similarity to SARS-CoV-2 than pMMoV.

## 4. Conclusion

By directly measuring N1, N2, BCoV, and pMMoV, we assessed the recovery and practicality of different concentration methods required for SARS-CoV-2 WBE. HA filtration with bead beating showed high recovery of all gene targets at a significant distance above the LoQ, leading to low variability across measurements. The same method also demonstrated relatively moderate startup costs, low cost per sample, and high throughput. HA filtration with bead beating is therefore a preferred concentration method in many situations from the perspective of recovery and practicality. Additional attention should be paid optimizing the sensitivity and recovery of all concentration methods. Future work should also identify the form of SARS-CoV-2 RNA and establish how this affects measurements via the different concentration methods. Additionally, more work needs to be done to determine the best process controls and normalization factors for SARS-CoV-2. The WBE for SARS-CoV-2 community needs to identify process controls and surrogates that behave similarly to SARS-CoV-2 in wastewater and Overall, this work further demonstrates that methods to concentrate SARS-CoV-2 RNA for WBE are low cost and reliable.

## Data Availability

Data can be found in the body of the manuscript itself or in the supplementary information.

## Acknowledgements

We thank Kathy Ensor for her invaluable contributions to the implementation of a wastewater monitoring system in Houston. We thank Paul Zappi, Rae Mills, Carol LaBreche, Walid Samarneh, and Aisha Niang from Houston Water for their assistance in collecting wastewater samples. We thank Lilian Mojica, Braulio Garcia, Courtney Hundley, Jeremy Rangel, Kelsey Caton, Rebeca Schneider, Daniel Bahrt, Kaavya Damakonda, Patrick Key, and Naomi Macias from the Houston Health Department for their assistance in sample collection and data analysis. We thank Kristina Cibor, Esther Lou, Basmah Maiga, Camille McCall, and Pavan Raja, and for their assistance in sample collection, processing, analysis, and project management. We thank Jeseth Delgado Vela, Adam Smith, Nadine Kotlarz, Francis de los Reyes, and Angela Harris for their discussions on wastewater-based epidemiology.

---

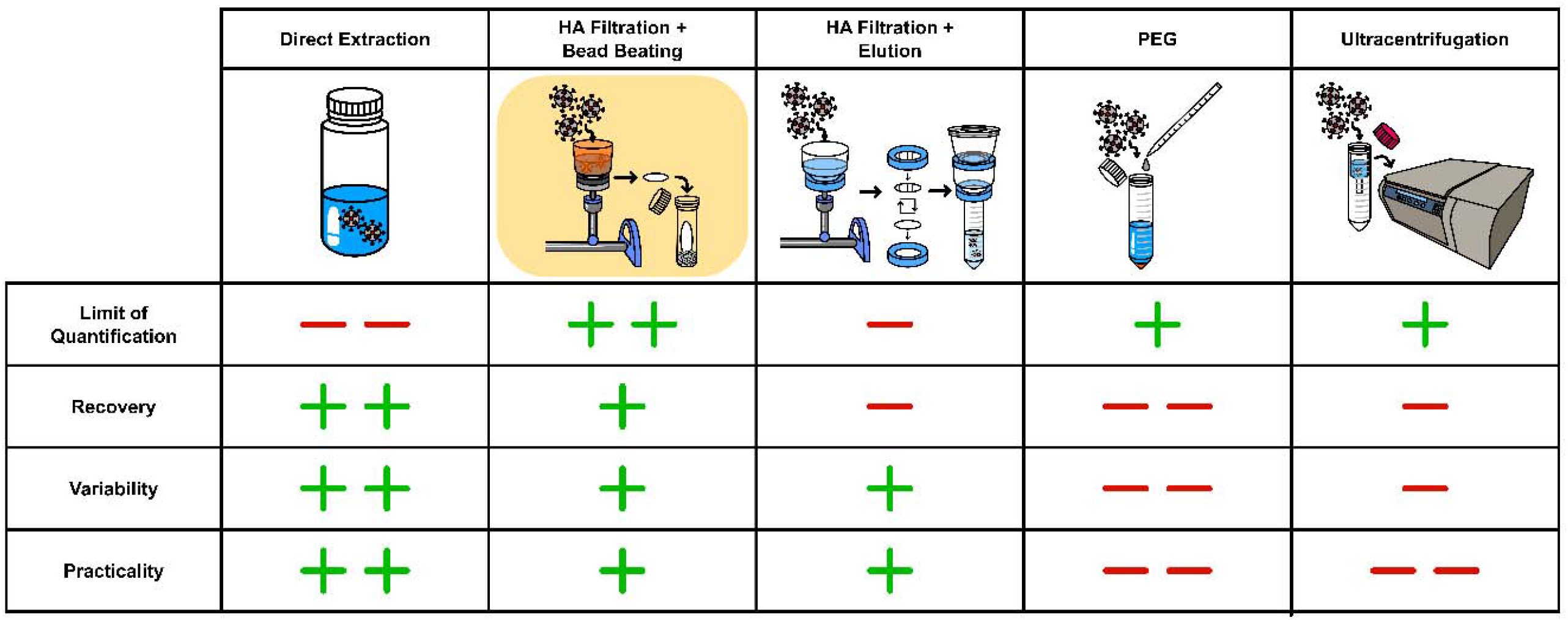
Estimates of the relative relationship of the five concentration methods to each other based on different performance characteristics.

