## Supplemental Information for "Evaluating recovery, cost, and throughput of different concentration methods for SARS-CoV-2 wastewater-based epidemiology"

**This file includes:**

*Quantification Method Information* **-** Table S1 to S7 (page 2-4)

*Concentration Factor Calculations* - Table S8 to S13 (page 5-6)

*Startup Cost Calculations -* Table S14 to S18 (page 7-8)

*Consumable Cost Calculations -* Table S19 to S22 (page 9-10)

*Throughput Analysis* - Table S23 (page 11)

Quantification Method Information

**Table S1.** Primers and probes used for quantification of SARS-CoV-2, BCoV, and pMMoV.

| **Quantification** **Method** | **Assay Name** | **Sequence (5’ - 3’)** |
| --- | --- | --- |
| RT-ddPCR | SARS-CoV-2 CDC N1 | **Fwd** - GACCCCAAAATCAGCGAAAT  **Rev** - TCTGGTTACTGCCAGTTGAATCTG  **Probe** - HEX - ACCCCGCATTACGTTTGGTGGACC - BHQ-1 |
| RT-ddPCR | SARS-CoV-2 CDC N2 | **Fwd** - TTACAAACATTGGCCGCAAA  **Rev** - GCGCGACATTCCGAAGAA  **Probe** - FAM - ACAATTTGCCCCCAGCGCTTCAG - Zen, Iowa Black FQ |
| RT-ddPCR | BCoV_M | **Fwd** - CCAGCTTATGTGACTGTTGCT  **Rev** - GCAAAACCACTAGTATCGCCT  **Probe** - FAM - TCTCACACCTGCTCACGTATAAGCG - QSY |
| RT-qPCR | pMMoV_VGP2 | **Fwd** - GAGTGGTTTGACCTTAACGTTTGA  **Rev** - TTGTCGGTTGCAATGCAAGT  **Probe** - JUN - CCTACCGAAGCAAATG - QSY |

Table S2. Reaction mixture composition for ddPCR of N1/N2.

| **Reagent** | **Volume (µL)** |
| --- | --- |
| H2O | 0.12 |
| Supermix | 5.5 |
| Reverse Transcriptase | 2.2 |
| 300 mM DTT | 1.1 |
| N1 Primer/Probe Mix | 1.1 |
| N2 Primer/Probe Mix | 1.1 |
| Template | 10 |

**Table S3.** Reaction mixture composition for ddPCR of BCoV..

| **Reagent** | **Volume (µL)** |
| --- | --- |
| H2O | 8.1 |
| Supermix | 5.5 |
| Reverse Transcriptase | 2.2 |
| 300 mM DTT | 1.1 |
| BCoV Primer/Probe Mix | 1.1 |
| Template (1 pg – .5 µg) | 10 |

**Table S4.** Reaction mixture composition for qPCR of pMMoV.

| **Reagent** | **Volume (µL)** |
| --- | --- |
| H2O | 0.4 |
| 2x qPCRBIO Probe 1-Step Go No-ROX | 5.0 |
| 20x RTase Go | 0.1 |
| pMMoV Primer/Probe Mix | 0.5 |
| Template (1 pg – .5 µg) | 4.0 |

**Table S5.** Composition of primer/probe mixes used in ddPCR and qPCR. Concentrations in the table are 20x of the final concentration in the reaction mix.

| **Component** | **ddPCR (N1, N2, BCoV)*** | **qPCR (pMMoV)*** |
| --- | --- | --- |
| Probe | 5 µM | 4 µM |
| Forward Primer | 18 µM | 8 µM |
| Reverse Primer | 18 µM | 8 µM |

**Table S6.** Thermocycling conditions for ddPCR of N1/N2 and BCoV.

| **Step** | **Temperature** | **Time** | **Cycles** |
| --- | --- | --- | --- |
| Reverse Transcription | 50 °C | 60 minutes | N/A |
| Enzyme Activation | 95 °C | 10 minutes |  |
| Denaturation | 95 °C | 30 seconds | 40x (Ramp rate = 2 °C/sec) |
| Annealing, Extension | 60 °C | 1 minute |  |
| Enzyme Deactivation | 98 °C | 10 minutes | N/A |
| Hold | 4 °C | Inf. |  |

**Table S7.** Thermocycling conditions for ddPCR of N1/N2 and BCoV.

| **Step** | **Temperature** | **Time** | **Cycles** |
| --- | --- | --- | --- |
| Reverse Transcription | 50 °C | 5 minutes | N/A |
| Enzyme Activation | 95 °C | 20 seconds |  |
| Denaturation | 95 °C | 3 seconds | 40x |
| Annealing, Extension | 60 °C | 30 seconds |  |

Concentration Factor Calculations

| **Table S8. Direct extraction** | | | |
| --- | --- | --- | --- |
|  | **Volume** |  | **Concentration Factor** |
| Starting Volume: | 300 | ul |  |
| Elution Volume: | 100 | ul | 3 |
|  |  | **Total:** | 3 |

| **Table S9. HA filtration with bead beating** | | | |
| --- | --- | --- | --- |
|  | **Volume** |  | **Concentration Factor** |
| Starting Volume: | 50 | ml |  |
| Lysis buffer Added: | 700 | ul | 71.43 |
| Added to Chemagic: | 350 | ul |  |
| Elution Volume: | 100 | ul | 3.5 |
|  |  | **Total:** | 250 |

| **Table S10. HA filtration with elution** | | | |
| --- | --- | --- | --- |
|  | **Volume** |  | **Concentration Factor** |
| Starting Volume: | 25 | ml |  |
| NaOH Eluent: | 2.5 | ml | 10 |
| Eluent Volume: | 300 | ul |  |
| Elution Volume: | 100 | ul | 3 |
|  |  | **Total:** | 30 |

| **Table S11. PEG** |  |  |  |
| --- | --- | --- | --- |
|  | **Volume** |  | **Concentration Factor** |
| Starting Volume: | 200 | mL |  |
| Resuspension Volume | 2 | mL | 100 |
| Concentrate Volume: | 300 | ul |  |
| Elution Volume: | 100 | ul | 3 |
|  |  | **Total:** | 300 |

| **Table S12. Ultrafiltration** | | | |
| --- | --- | --- | --- |
|  | **Volume** |  | **Concentration Factor** |
| Starting Volume: | 50 | ml |  |
| Concentrate: | 1.5 | mL | 33.33 |
| Concentrate Volume: | 300 | ul |  |
| Elution Volume: | 100 | ul | 3 |
|  |  | **Total:** | 100 |

**Table S13.** Summary of concentration factors for each method.

| **Method** | **Concentration Factor** |
| --- | --- |
| Direct extraction | 3 |
| HA filtration with bead beating | 250 |
| HA filtration with elution | 30 |
| PEG | 300 |
| Ultrafiltration | 100 |

Startup Cost Calculations

| **Table S14. Direct extraction** | | | |
| --- | --- | --- | --- |
|  | **Cost** | **Manufacturer** | **Catalog Number** |
| Microcentrifuge | $5,650.00 | Thermo Scientific | 75-772-446 |
| **Total:** | **$5,650.00** |  |  |

| **Table S15. HA filtration with bead beating** | | | |
| --- | --- | --- | --- |
|  | **Cost** | **Manufacturer** | **Catalog Number** |
| Centrifuge | $9,000 | Thermo Scientific | 75-004-525 |
| Filter Manifold | $1,347.84 | Sterlitech | 180600-01 |
| Filter Holder | $253.38 | Sterlitech | 200300-01 |
| Vacuum Pump | $466.00 | Sterlitech | 167800 |
| Vacuum Flask | $59.12 | MilliporeSigma | XX1014705 |
| Graduated Cylinder | $33.84 | Thermo Scientific | 08-572-5D |
| Bead Beater | $4,208 | BioSpec | 112011 |
| **Total:** | **$15,368** |  |  |

| **Table S16. HA filtration with elution** | | | |
| --- | --- | --- | --- |
|  | **Cost** | **Manufacturer** | **Catalog Number** |
| Centrifuge | $9,000 | Thermo Scientific | 75-004-525 |
| Filter Manifold | $1,347.84 | Sterlitech | 180600-01 |
| Filter Holder | $253.38 | Sterlitech | 200300-01 |
| Vacuum Pump | $466.00 | Sterlitech | 167800 |
| Vacuum Flask | $59.12 | MilliporeSigma | XX1014705 |
| Graduated Cylinder | $33.84 | Thermo Scientific | 08-572-5D |
| **Total:** | **$11,160** |  |  |

| **Table S17. PEG** |  |  |  |
| --- | --- | --- | --- |
|  | **Cost** | **Manufacturer** | **Catalog Number** |
| Standing Centrifuge | $20,000.00 | Thermo Scientific | 75-006-580 |
| 500 mL Centrifuge Bottles | $288.13 | VWR | 47735-696 |
| **Total:** | **$20,288** |  |  |

| **Table S18. Ultrafiltration** | | | |
| --- | --- | --- | --- |
|  | **Cost** | **Manufacturer** | **Catalog Number** |
| Centrifuge | $9,000 | Thermo Scientific | 75-004-525 |
| **Total:** | **$9,000** |  |  |

Consumables Cost Calculations

| **Table S19. Direct extraction** | | | |  |  |
| --- | --- | --- | --- | --- | --- |
|  | **Cost per Package** | **Samples per Package** | **Cost per Sample** | **Manufacturer** | **Catalog Number** |
| 1.5 mL Centrifuge Tube | $68.30 | 500 | $0.14 | Eppendorf | 05-402-25 |
|  |  | **Total:** | **$0.14** |  |  |

| **Table S20. HA filtration with bead beating** | | | | |  |
| --- | --- | --- | --- | --- | --- |
|  | **Cost per Package** | **Samples per Package** | **Cost per Sample** | **Manufacturer** | **Catalog Number** |
| MgCl_2_ · 6H_2_O | $232.00 | 1967.5 | $0.12 | Sigma-Aldrich | M0250-500G |
| Filters | $340.00 | 600 | $0.57 | MilliporeSigma | HAWG047S6 |
| 50 mL Conical Tubes | $210.33 | 500 | $0.42 | Thomas Scientific | 1184R09 |
| 0.1 mm dia beads | $38.00 | 150 | $0.25 | BioSpec | 11079101 |
| Bead Beating Tubes | $70.50 | 500 | $0.14 | Fisherbrand | 02-682-558 |
|  |  | **Total:** | **$1.50** |  |  |

| **Table S21. HA filtration with elution** | | | |  |  |
| --- | --- | --- | --- | --- | --- |
|  | **Cost per Package** | **Samples per Package** | **Cost per Sample** | **Manufacturer** | **Catalog Number** |
| MgCl_2_, anhydrous | $232.00 | 1967.5 | $0.12 | MilliporeSigma | M0250-500G |
| 15 mL Conical Tubes | $193.06 | 500 | $0.39 | Thomas Scientific | 1184R08 |
| EZ-Fit™ Filtration Unit | $254.00 | 48 | $5.29 | MilliporeSigma | EFHAW100B |
| H_2_SO_4_ | $189.80 | 734154.5 | $0.0003 | Fisher Chemical | A300-212 |
| NaOH | $66 | 500037.5 | $0.0001 | Fisher Chemical | S318-100 |
|  |  | **Total:** | **$5.80** |  |  |

| **Table S22. PEG** |  |  |  |  |  |
| --- | --- | --- | --- | --- | --- |
|  | **Cost per Package** | **Samples per Package** | **Cost per Sample** | **Manufacturer** | **Catalog Number** |
| Bottle Top Filters | $120.00 | 12 | $10.00 | MilliporeSigma | SCGPS05RE |
| PEG 8000 | $46.78 | 62.5 | $0.75 | VWR | VWRV0159-1KG |
| NaCl | $93.00 | 342.2 | $0.27 | Fisher Chemical | S271-1 |
| PBS solution | $0.72 | 500 | $0.001 | Fisher Chemical | S375-500, S369-500, BP358-10 |
|  |  | **Total:** | **$11.02** |  |  |

| **Table S23. Ultrafiltration** | | | | | |
| --- | --- | --- | --- | --- | --- |
|  | **Cost per Package** | **Samples per Package** | **Cost per Sample** | **Manufacturer** | **Catalog Number** |
| 50 mL Conical Tubes | $210.33 | 500 | $0.42 | Thomas Scientific | 1184R09 |
| Amicon® filters | $277.00 | 24 | $11.54 | MilliporeSigma | ACS510024 |
| 1.5 mL Centrifuge Tube | $68.30 | 500 | $0.14 | Eppendorf | 05-402-25 |
|  |  | **Total:** | **$12.10** |  |  |

Throughput Analysis

**Table S23.** Breakdown of times in throughput analysis.

|  | **Time Required per 6 Samples (min)** | | | | | | | | | | |
| --- | --- | --- | --- | --- | --- | --- | --- | --- | --- | --- | --- |
|  | **Bead beating Tube Preparation** | **Sample and Manifold Preparation** | **Solid Removal Centrifugation** | **Centrifugation** | **Wait After MgCl_2_ Addition** | **Filtration** | **Elution** | **Precipitation** | **Sample Storing** | **Total** | **Total (hr)** |
| **HA filtration with bead beating** | 6 | 5 | 10 | 0 | 5 | 15 | 0 | 0 | 2 | 43 | 0.72 |
| **HA filtration with elution** | 0 | 5 | 10 | 0 | 5 | 3 | 6 | 0 | 2 | 31 | 0.52 |
| **Ultrafiltration** | 0 | 10 | 10 | 70 | 0 | 0 | 0 | 0 | 2 | 92 | 1.53 |
| **Direct Extraction** | 0 | 0 | 5 | 0 | 0 | 0 | 0 | 0 | 2 | 7 | 0.12 |
| **PEG** | 0 | 0 | 15 | 0 | 0 | 20 | 0 | 240 | 2 | 277 | 4.62 |
